# A precise measure of the impact of the first wave of Covid-19 on life expectancy. Regional differentials in Switzerland

**DOI:** 10.1101/2020.11.19.20234716

**Authors:** Philippe Wanner

## Abstract

Based on publicly available data supplied by the Swiss Federal Statistical Office (FSO), we calculated life tables by sex and by week for seven major regions of Switzerland in 2020, up to October 26th. These life tables provide information on the trends of life expectancy at birth and at the age of 65 years during the first wave of the coronavirus disease 2019 (COVID-19) epidemic.

The results show a strong cyclical decrease in life expectancy, particularly in Ticino, where this variable has decreased by almost 6 years compared to the 2019 life expectancy, and in the Lake Geneva region. The other regions of Switzerland observed more modest decreases during the first wave, generally not exceeding a 2-year reduction. This decrease can be explained to some extent by seasonal variations in this indicator.

In conclusion, the very sharp decrease in the average lifespan observed in the two regions mentioned above suggests that the first wave of the epidemic had a significant impact. It also reflects an unfavourable health situation. The life expectancy at the age of 65 years observed at the end of March 2020 in Ticino corresponded to the average life expectancy observed in Switzerland forty years ago.

The calculated indicators have the advantage of accounting for the age structures of the respective populations. They therefore demonstrate their usefulness in monitoring during a pandemic, such as the one occurring currently.

## Introduction

The first wave of coronavirus disease 2019 (COVID-19) in March and April 2020 caused significant excess mortality in almost all European countries. In Switzerland, this excess mortality has been documented by raw data from the Federal Statistical Office (FSO) ; the data indicate a higher number of deaths in those aged 65 and over than the average number of deaths in the same population in previous years. In contrast, before the age of 65, the mortality rate remained more or less the same. This excess mortality was short-lived, lasting from Mid-March to April.

The increase in the number of deaths, compared to previous years, is not an uncommon phenomenon. It has been documented during various influenza episodes. However, this increase in the level of mortality is difficult to comment on without a more accurate and age-adjusted measure of mortality.

For this reason, the establishment of life tables and the calculation of life expectancies can provide an indication of the precise impact of an epidemic on the population as a whole. Indeed, life expectancies are standardized indicators that reflect the rate of mortality in a given period after taking into account the age structure of the population and the distributions of deaths by age. They also allow spatial comparisons of health situations. In our case, they can indicate the net impact of a public health issue on the life span of a population.

In this paper, we calculate life expectancies in seven major regions of Switzerland and describe the short-term consequences of the first wave of COVID-19 on life expectancy.

## Methods

Life tables were calculated from data on the number of deaths published by the FSO for the different regions of Switzerland. We stratified the number of deaths (all-cause death) by five-year age groups, region of Switzerland, sex and week for the first 45 weeks of 2020. The populations by sex and age as of December 31, 2019, were used as the basis for the calculation of mortality rates in each age group, which were transformed into quotients according to the Berkson formula. In order to have a reference value, we also computed the weekly and annual life tables for the year 2019.

To do so, we assumed a uniform distribution of deaths over the considered age groups and a life expectancy of 2.5 years in the last open age group. Since the 2020 population by age and sex in the target regions of Switzerland is unknown, the rates were therefore calculated by dividing deaths by the population at the end of December 2019. However, the populations in the different regions are likely to change due to births, deaths, and migration flow. Given the short period under consideration, changes in populations should not significantly influence the results.

Based on these data, we calculated weekly life expectancies at different ages in seven major regions of Switzerland and by sex. Due to the low number of weekly deaths in the smaller regions, statistical fluctuations may be present. For this reason, we smoothed the weekly values by applying a moving average, which was calculated by considering the average value of three weeks. Only life expectancies at birth and at age 65 are presented here.

## Results

Table 1 shows the minimum life expectancy during the first wave (March 23 to April 16) compared with the 2019 annual average. According to this table, two regions were significantly affected by this first wave. First, considering the average over the whole period, life expectancy at birth in Ticino decreased by 6 years in men and women; it was during the week of March 23 for men and March 30 for women that Ticino life expectancies were the lowest, at 76.8 years in men and 79.7 years in women. At the age of 65, male life expectancy was less than 14 years (female 16 years), which is more than 6 years less than the reference period (2019).

**Table 1:** Average Life Expectancy in 2019 and Life Expectancy during the 1^st^ Wave, according to Sex, age and Region.

|  | Arc<br>lémanique | Ticino | Espace<br>Mitteland | Nordwest<br>-schweiz | Zurich | Ostschweiz | Zentralschweiz |
| --- | --- | --- | --- | --- | --- | --- | --- |
| Men |  |  |  |  |  |  |  |
| E0 |  |  |  |  |  |  |  |
| **Gap** | **3.70** | **5.75** | **2.16** | **1.86** | **2.03** | **1.94** | **1.86** |
| Average 2019 | 81.64 | 82.59 | 81.38 | 81.76 | 81.77 | 81.20 | 82.08 |
| 1st wave | 77.94 | 76.84 | 79.22 | 79.90 | 79.74 | 79.26 | 80.22 |
| E65 |  |  |  |  |  |  |  |
| **Gap** | **3.89** | **6.21** | **1.72** | **1.98** | **1.98** | **1.57** | **1.39** |
| Average 2019 | 19.67 | 20.14 | 19.35 | 19.75 | 19.67 | 19.28 | 19.86 |
| 1st wave | 15.78 | 13.93 | 17.63 | 17.77 | 17.69 | 17.71 | 18.47 |
| Women |  |  |  |  |  |  |  |
| E0 |  |  |  |  |  |  |  |
| **Gap** | **2.16** | **5.93** | **0.99** | **0.80** | **1.69** | **1.56** | **0.44** |
| Average 2019 | 85.25 | 85.61 | 84.65 | 84.65 | 84.73 | 84.69 | 85.05 |
| 1st wave | 83.09 | 79.68 | 83.66 | 83.85 | 83.04 | 83.12 | 84.61 |
| E65 |  |  |  |  |  |  |  |
| **Gap** | **2.02** | **6.39** | **0.69** | **1.20** | **0.92** | **0.54** | **0.75** |
| Average 2019 | 22.12 | 22.40 | 21.62 | 21.67 | 21.76 | 21.64 | 21.98 |
| 1st wave | 20.10 | 16.01 | 20.93 | 20.47 | 20.84 | 21.10 | 21.23 |

Computations are based on FSO data.

The Arc lémanique, for the most part, observed a reduction of more than 3.5 years in male life expectancy (at birth and at the age of 65) and nearly 2 years in female life expectancy. The lowest levels of life expectancy were observed in the week beginning on March 23 in men and on March 30 in women.

The other regions of Switzerland experienced shorter life expectancies during the first wave than the annual averages. The reduction in life expectancy at birth was approximately 2 years in men and between 0.8 years and 1.9 years in women. Similar values were observed for life expectancy at age 65. These decreases in life expectancy indicate a small impact of mortality associated with the first wave; however, the decrease is hardly more than that associated with normal seasonal variations.

Annex 1 provides the trends in the life expectancy at birth and at age 65 for every region and for both sexes. The data refers to the period until week 45 (2-8 November). Undoubtedly, life expectancies in some regions that were spared during the first wave are declining rapidly since the end of October.

## Conclusion

In times of health crises, it is important to have reliable data to monitor the precise health situation. The availability of mortality data is one of the elements of COVID-19 monitoring. These data have highlighted excess mortality during successive waves compared to the number of expected deaths on the basis of recent years in many countries and regions.

The calculation of life expectancies provides precise information, as this indicator controls for the age structures of populations. It allows comparisons on a standardized basis. It measures the impact of a health crisis in terms of the length of remaining life for the entire population (life expectancy at birth) or for elderly individuals (life expectancy at age 65) given a specific health-based context. The results obtained show the different impacts of the first wave in each region. They showed a very sharp drop in life expectancy, indicating significant health impairment that was not limited to elderly individuals.

The situation in Ticino at the end of March is particularly remarkable. Men aged 65 and over had a level of mortality equivalent to a life expectancy of 14 years (16 years for women). Such values were last exceeded for all of Switzerland in 1980. In other words, the health situation in Ticino during the first wave was comparable to that observed 40 years ago.

Until now, two papers measured the impact of Covid19 on life expectancy. Gishlandi et al., 2020 showed a significant decrease in life expectancy in five provinces of Northern Italy and found that for the first wave of the epidemic, life expectancy at birth reduced by 5.1 to 7.8 for men and 3.2 to 5.8 years for women. Our results from Ticino are therefore comparable to those observed in neighbouring Lombardy. Trias-Llimos et al., 2020 for Spain showed that the drop in weekly life expectancy reached 7.6 years and at the national level and 15 years in Madrid.

Calculations may soon be performed for the second wave, which occurred beyond the period covered by the currently available data. The approach will thus make it possible to compare the respective impacts of the two waves using a single indicator.

## Data Availability

Data that are used are available to the website of the Swiss Federal Statistical office (www.bfs.admin.ch). Data being subject to weekly revisions, a copy of the data downloaded on November 11, 2020 is available from the author at request

## Annexe

### Life expectancy at birth and at the age of 65 according to the Swiss region and the sex

#### Ticino

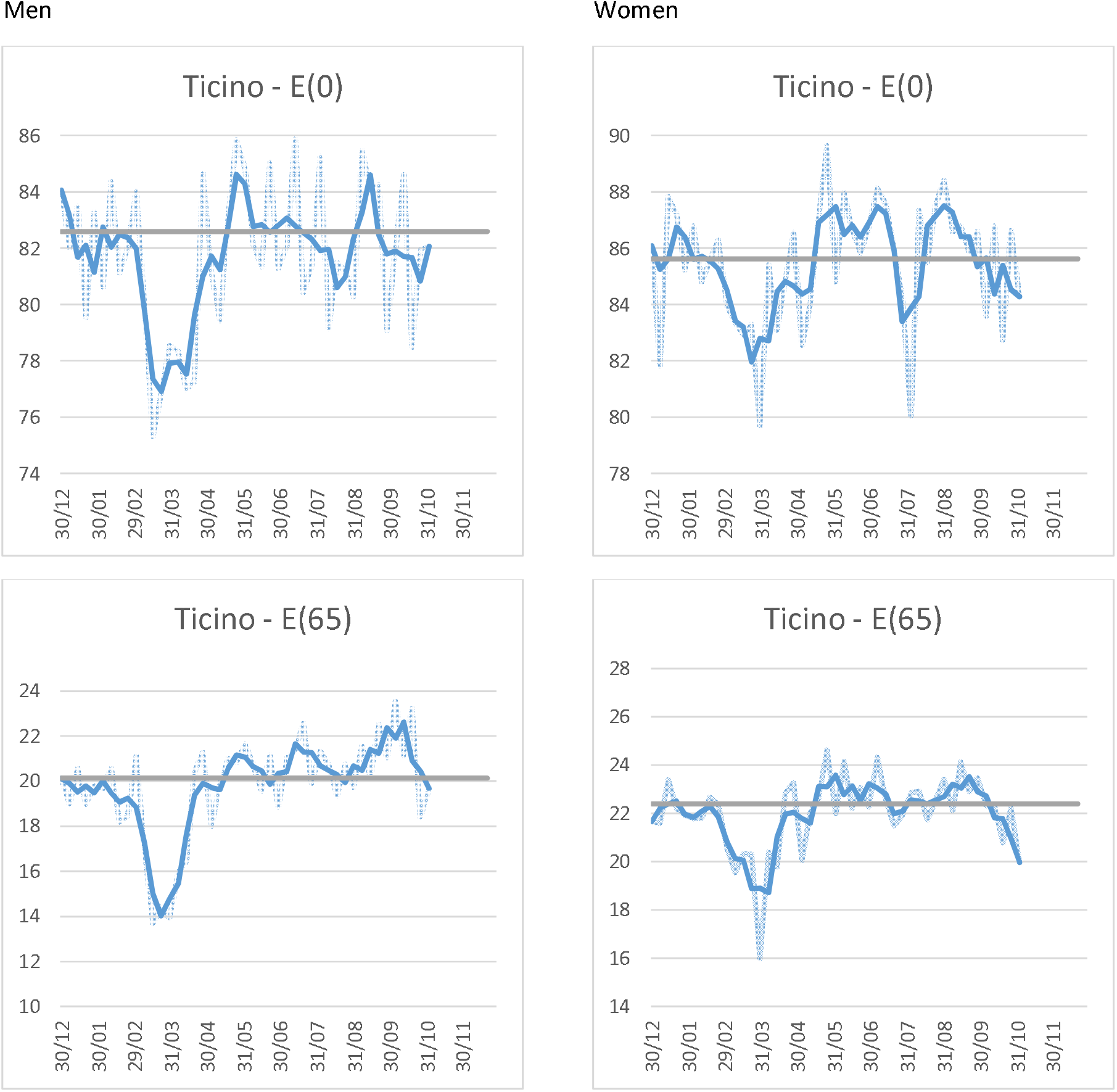

The light blue represents the weekly values, the dark blue the smoothed values. They grey line represents the life expectancy for the year 2019. E(0) = Life expectancy at birth; E(65) = Life expectancy at the age of 65.

Life expectancies are based on the FSO data by region, age group (5 year intervals) and gender. They are computed based on the permanent resident population at the 31.12.2019.

### Arc Lémanique

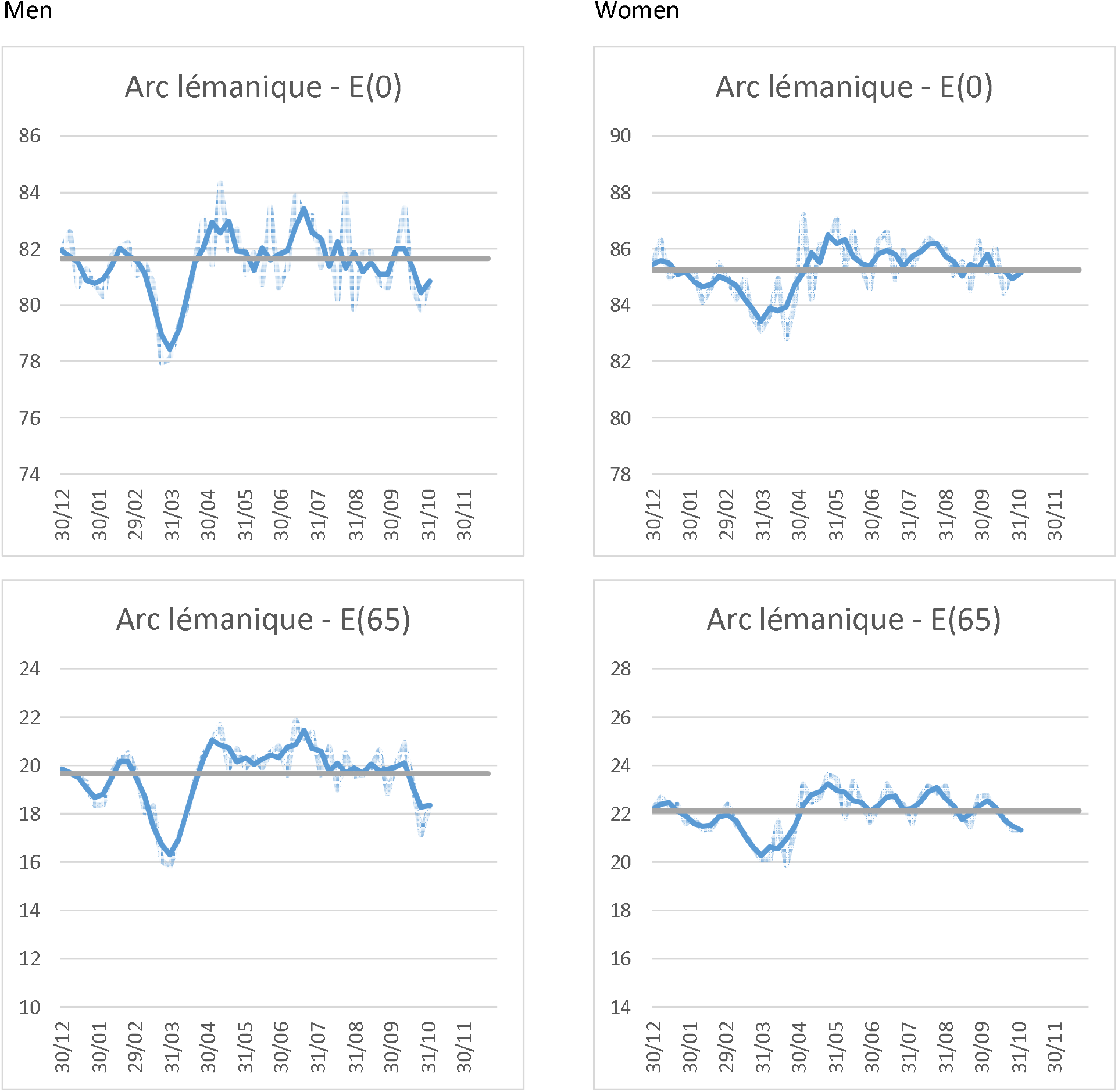

The light blue represents the weekly values, the dark blue the smoothed values. They grey line represents the life expectancy for the year 2019. E(0) = Life expectancy at birth; E(65) = Life expectancy at the age of 65.

Life expectancies are based on the FSO data by region, age group (5 year intervals) and gender. They are computed based on the permanent resident population at the 31.12.2019.

### Zürich

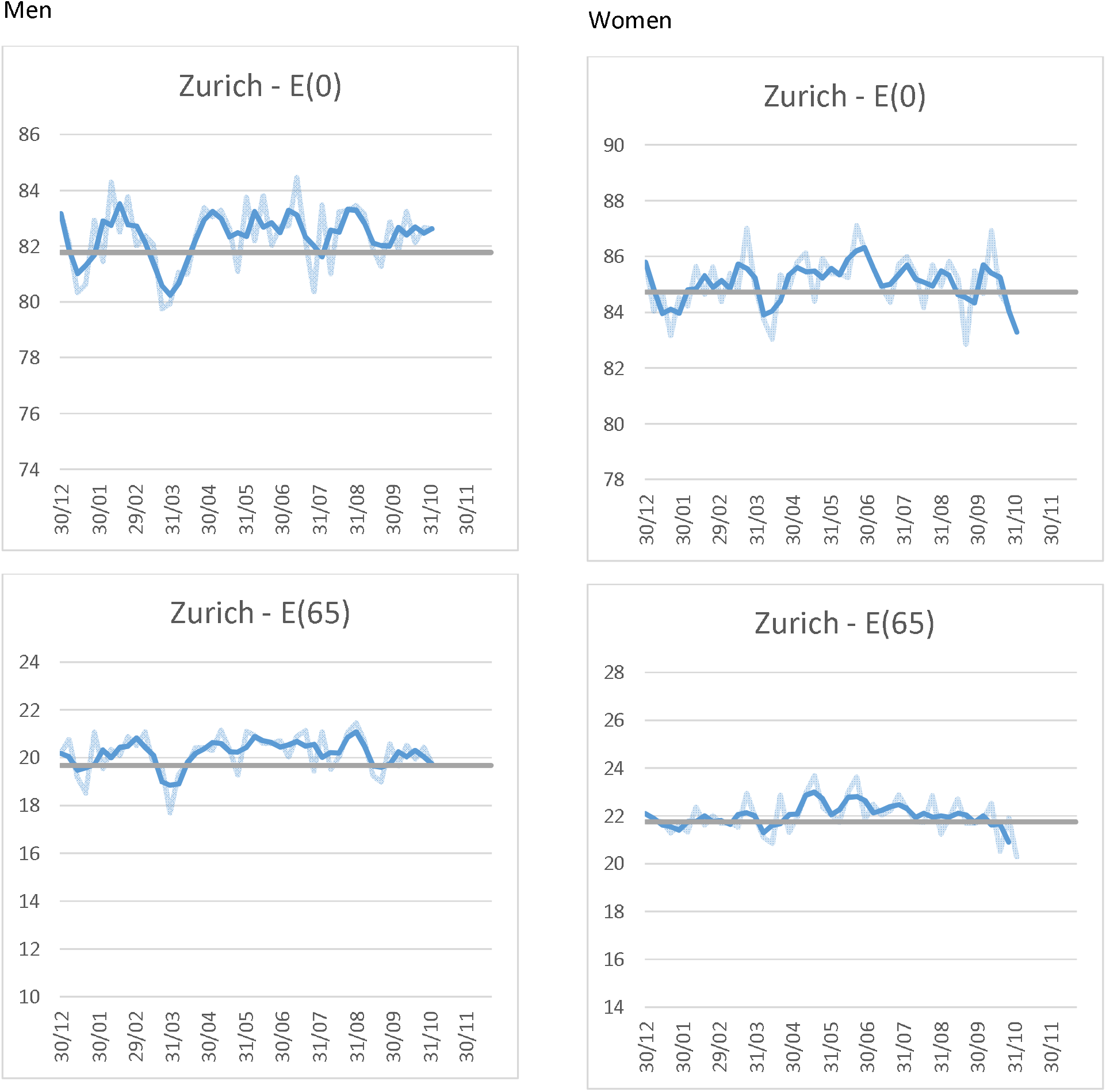

The light blue represents the weekly values, the dark blue the smoothed values. They grey line represents the life expectancy for the year 2019. E(0) = Life expectancy at birth; E(65) = Life expectancy at the age of 65.

Life expectancies are based on the FSO data by region, age group (5 year intervals) and gender. They are computed based on the permanent resident population at the 31.12.2019.

### Espace Mitteland

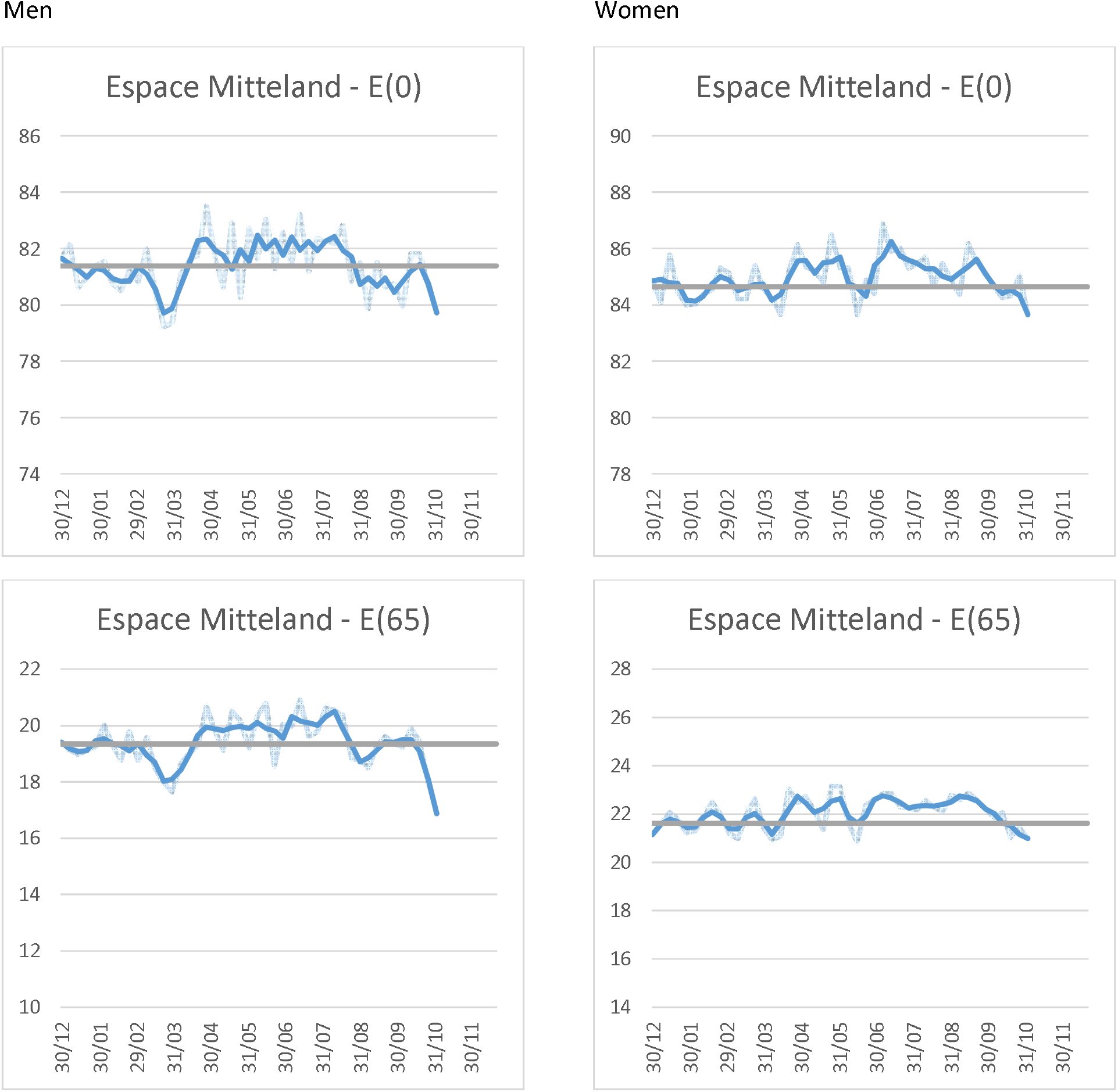

The light blue represents the weekly values, the dark blue the smoothed values. They grey line represents the life expectancy for the year 2019. E(0) = Life expectancy at birth; E(65) = Life expectancy at the age of 65.

Life expectancies are based on the FSO data by region, age group (5 year intervals) and gender. They are computed based on the permanent resident population at the 31.12.2019.

### Nordwestschweiz

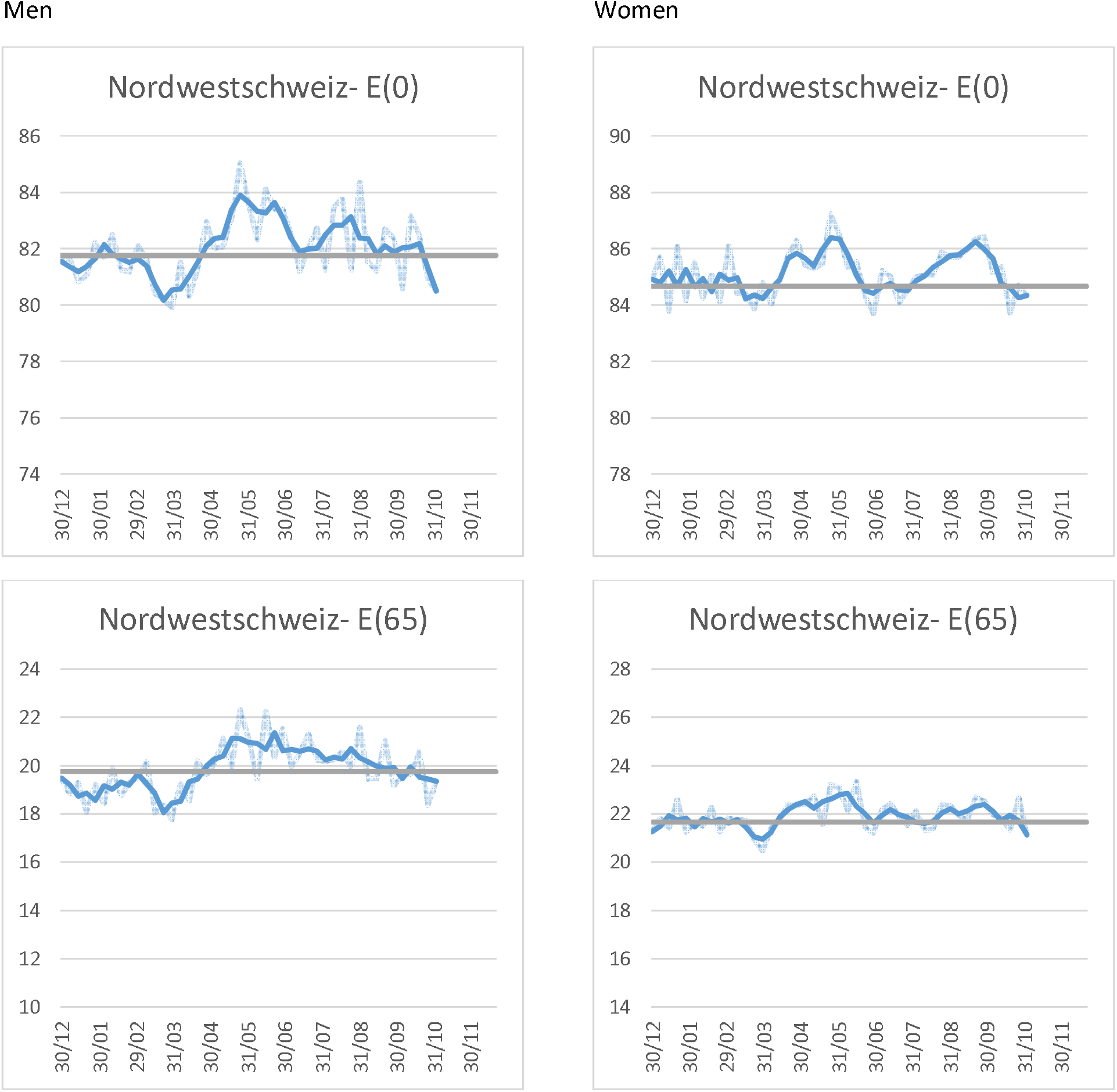

The light blue represents the weekly values, the dark blue the smoothed values. They grey line represents the life expectancy for the year 2019. E(0) = Life expectancy at birth; E(65) = Life expectancy at the age of 65.

Life expectancies are based on the FSO data by region, age group (5 year intervals) and gender. They are computed based on the permanent resident population at the 31.12.2019.

### Ostschweiz

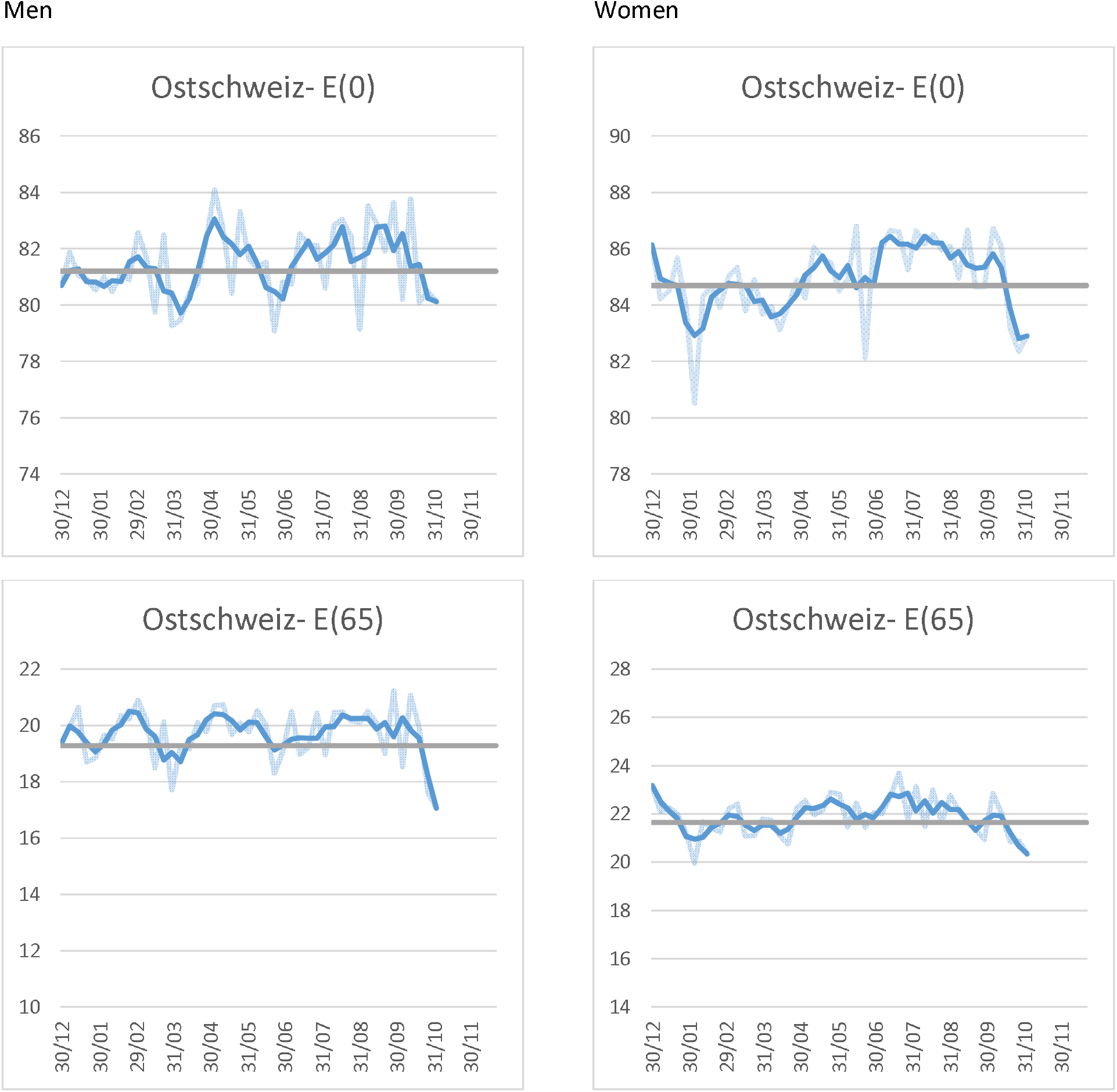

The light blue represents the weekly values, the dark blue the smoothed values. They grey line represents the life expectancy for the year 2019. E(0) = Life expectancy at birth; E(65) = Life expectancy at the age of 65.

Life expectancies are based on the FSO data by region, age group (5 year intervals) and gender. They are computed based on the permanent resident population at the 31.12.2019.

### Zentralschweiz

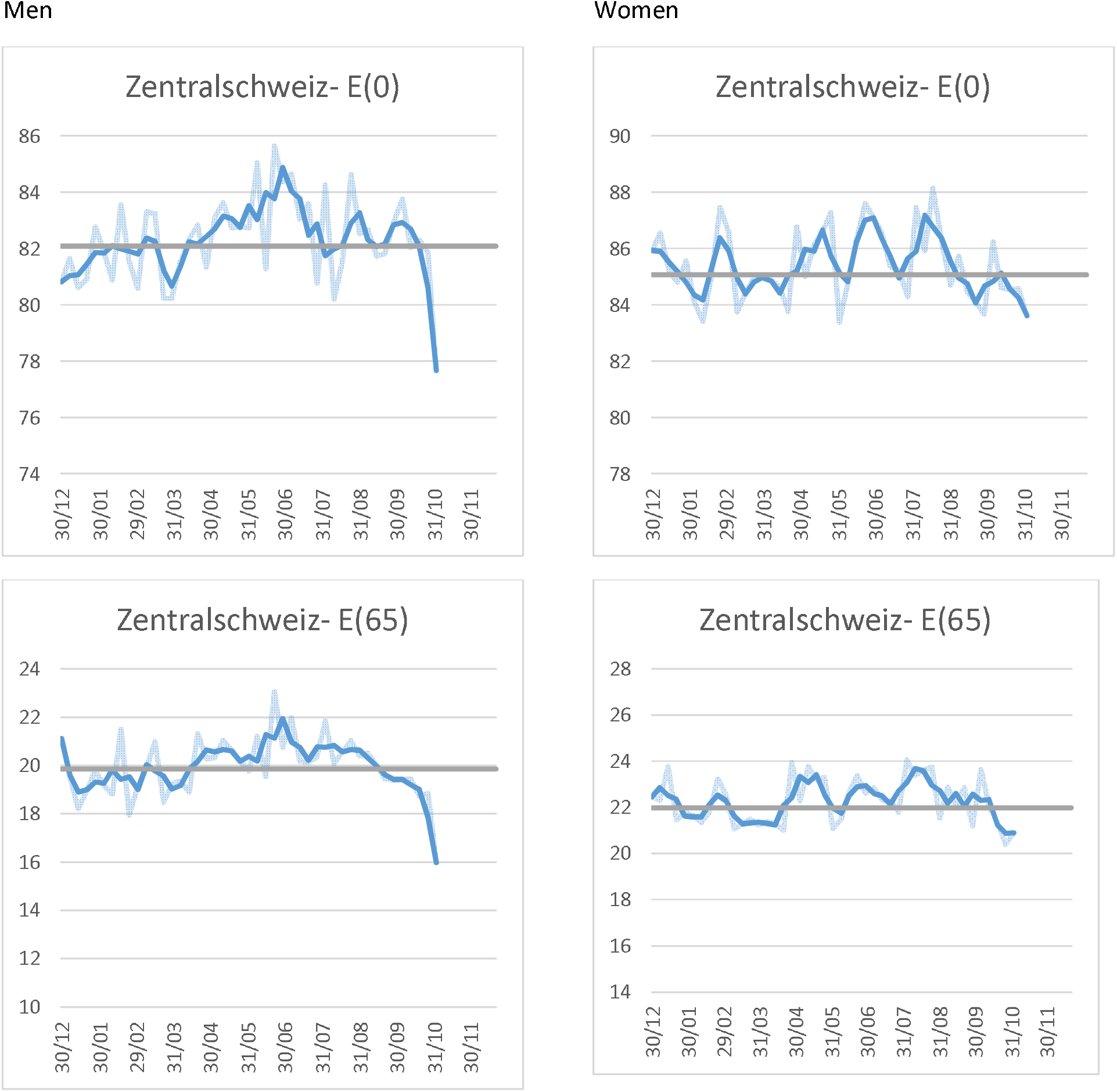

The light blue represents the weekly values, the dark blue the smoothed values. They grey line represents the life expectancy for the year 2019. E(0) = Life expectancy at birth; E(65) = Life expectancy at the age of 65.

Life expectancies are based on the FSO data by region, age group (5 year intervals) and gender. They are computed based on the permanent resident population at the 31.12.2019.

### Footnotes

1 Weekly number of deaths. https://www.bfs.admin.ch/bfs/en/home/statistics/health/state-health/mortality-causes-death.html Consulted on 18.11.2020.

